# Adapting for the COVID-19 pandemic in Ecuador, a characterization of hospital strategies and patients

**DOI:** 10.1101/2020.07.25.20161661

**Authors:** Daniel Garzon-Chavez, Daniel Romero-Alvarez, Marco Bonifaz, Juan Gaviria, Daniel Mero, Narcisa Gunsha, Asiris Perez, María Garcia, Hugo Espejo, Franklin Espinosa, Edison Ligña, Mauricio Espinel, Emmanuelle Quentin, Enrique Teran, Francisco Mora, Jorge. Reyes

## Abstract

The World Health Organization (WHO) declared coronavirus disease-2019 (COVID-19) a global pandemic on March 11^th^. In Ecuador, the first case of COVID-19 was recorded on February 29^th^. Despite efforts to control its spread, SARS-CoV-2 overrun the Ecuadorian public health system which became one of the most affected in Latin America with 22,719 cases reported up to April, 24^th^. Public health control measures, including social distancing and lockdowns, were implemented at different times in the biggest cities of the country: Guayaquil and Quito. The Hospital General del Sur de Quito (HGSQ) had to transition from a general to a specific COVID-19 health center in a short period of time to fulfill the health demand from patients with respiratory afflictions. Here, we summarized the implementations applied in the HGSQ to become a COVID-19 exclusive hospital, including the rearrangement of hospital rooms and a triage strategy based on a severity score calculated through an artificial intelligence (AI)-assisted chest computed tomography (CT). Moreover, we present clinical, epidemiological, and laboratory data from 75 laboratory tested COVID-19 patients, which represent the first outbreak of Quito city.

## Introduction

After the first announced cases of a pneumonia of unknown etiology in China in mid-December, 2019 [1], Around four million cases of COVID-19 have been reported worldwide for mid-May [2,3]. In South America, Ecuador occupies the second place in case numbers, just behind Brazil [3]. Ecuadorian implementations to control COVID-19 are relevant as a study case considering its fragmented public health system [4,5] and the heterogeneous evolution of the outbreak in its different administrative units (i.e., provinces). Two of its main cities, Quito and Guayaquil, applied recommended control measures, including social isolation, at different times. Guayaquil allowed mass gatherings and delayed strict isolation around two weeks in relation to Quito [6,7].

The first case in Ecuador was reported on February 29^th^, 2020 [8]. By April 7^th^, Ecuador managed diagnosis of SARS-Cov-2 mainly centralizing real-time reverse-transcriptase polymerase chain reaction testing (RT-PCR) of nasal/pharyngeal swabs in the National Institute of Public Health (INSPI, Spanish). As a consequence, reports were dependent on availability of resources and infrastructure; highly biasing official released case counts (Fig 1, [9]. For example, on April 10^th^, a release of 2,195 cases showed a sudden spike of COVID-19 in the country followed by reports of 97,209, and 63 cases the next three days (Fig 1, [9]). On April 24^th^, 11,000 positive cases of COVID-19 were released from one day to the other for a total of 22,719 positive cases, without clear justification on the sudden release; the majority of these cases were distributed on the most affected provinces: Guayas and Pichincha (Fig. 1; Supplementary_1, [9]). For April 23^th^, there were a total of 34,420 swab samples taken with results available for 23,383 (i.e., 67.93% tests completed), however, the sudden release of results on April 24^th^ was also accompanied with the report of 56,513 swab samples obtained (i.e., 22,093 in a single day); at this time, results were completed for 45,857 (i.e., 81.14%; Supplementary_1, [9]). This situation demonstrated an example of epidemiological misreporting problems and data ‘retention’ [10]. An initiative to decentralized testing led by University laboratories suggested in March 12^th^ had the potential to increase RT-PCR diagnosis availability [11], although for April 15^th^ 2020 it was still on plans of implementation [12].

**Figure 1.**
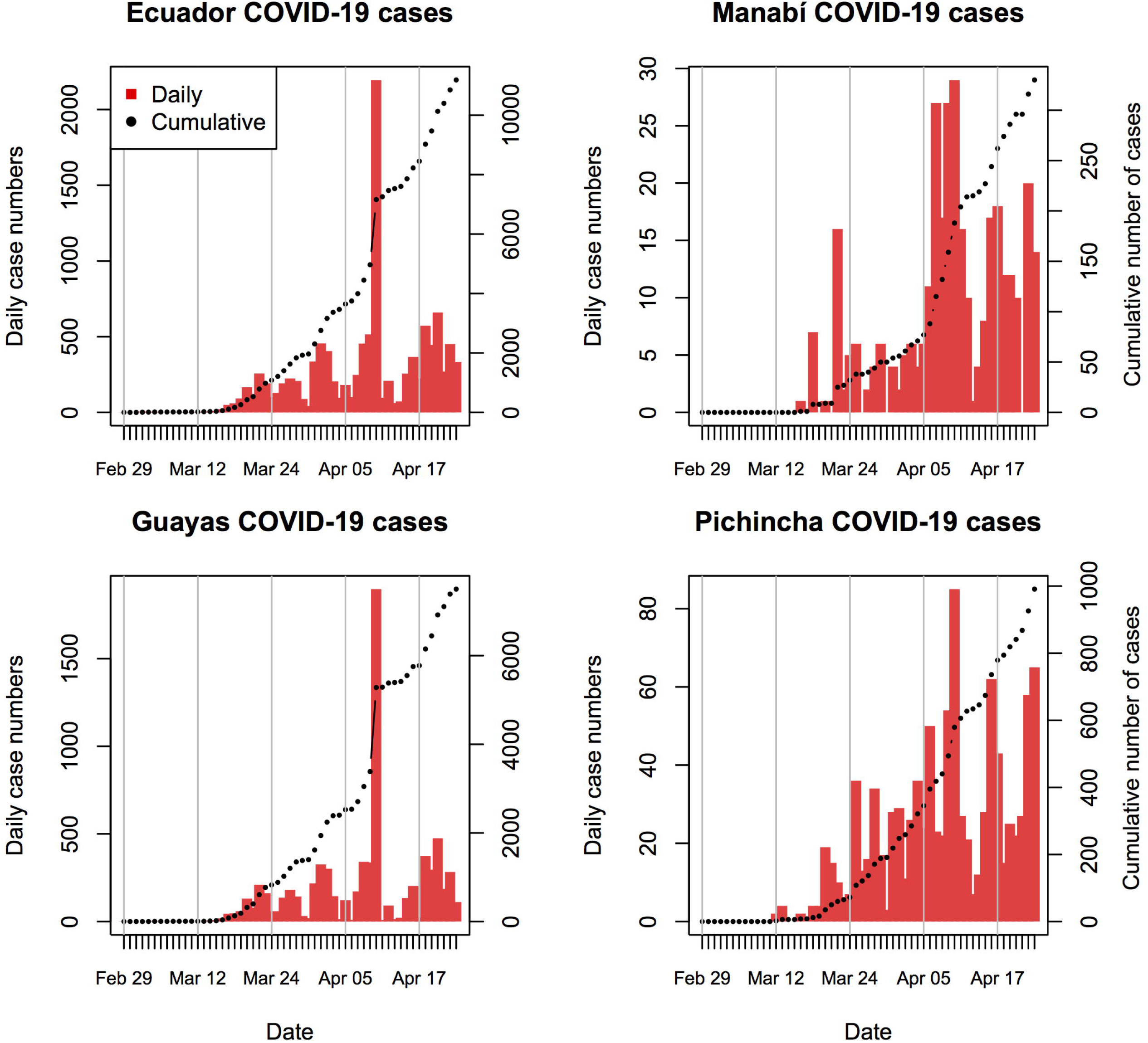
Official cumulative and daily case counts of COVID-19 in Ecuador up to April 23th, 2020. Plots are depicted for the whole country Ecuador (top-left) and the three most affected provinces: Manabí (top-right, for reference), Guayas (bottom-left), and Pichincha (bottom-right). We avoided plotting cases on April 24 because the addition of 11,000 cases in one day hinders the interpretation from the curves.

Due to the limitations on testing capacity, on-site approaches for patient triage have been suggested and actively explored using chest computed tomography (CT) or even pure clinical approaches [13,14]. Chest CT is performed to the majority of hospitalized patients with COVID-19 with main findings including the presence of ground glass opacities (GGO) [15–17]. At least 20% of non-severe COVID-19 infections have shown lack of changes in chest CT scans, while only 3% of severe patients present normal CTs [15]. Thus, the role of CT in severity screening and diagnosis is been evaluated thoroughly in different parts of the world and recommended as essential part of COVID-19 diagnosis in different guidelines [18–21].

The Hospital General del Sur de Quito (HGSQ, Spanish), was inaugurated on December 5^th^, 2017 as a center with 450 beds, providing secondary health-care with capabilities to solve third level health related complexities. It has medical specialties and subspecialties. The HGSQ provided medical care to an average of 20,000 monthly patients and attended 1,104 annual surgeries. On February 14^th^, the hospital was designated to become a COVID-19 specific treatment center. This nomination implied that suspected and diagnosed cases of COVID-19 were going to be attended exclusively by HGSQ personnel and therefore measures assuring a safe environment for patients and health workers had to be developed and implemented in a constrained schedule.

During the COVID-19 pandemic, the World Health Organization (WHO) has expressed concerns about health workers safety because of infections, burn out, stigmatization, and social reactions including physiological and physical violence [22]. Hospital infection rate in Wuhan was determined at around 4% and the number of health workers fatalities remained to be assessed [23]. A pandemic event represents a unique challenge for a hospital response which focuses mainly on preserving biosafety, avoiding nosocomial infections, coping with likely absenteeism of ill or death health personnel, and how to manage traditional diseases and chronic patients [24].

In the following manuscript, we describe the steps that drove the transition from a general to a COVID-19 exclusive hospital in a sanitary emergency background. First, we describe the utilization of an innovative triage approach based on CT-artificial intelligence (AI) in the context of limited availability of RT-PCR testing. Second, we describe the socio-demographic and clinical characteristics of the first 75 patients tested and treated for SARS-CoV-2 during the transition of this health facility. Finally, due to the high risk of asymptomatic carriers and super spreading events associated with coronavirus infections [17,25,26], we plotted the geographic distribution of 126 health care workers attending COVID-19 cases at the HGQS plus 54 laboratory tested COVID-19 cases, to potentially direct monitoring efforts in different areas of Quito city.

## Methods

### Ethic statement

This study was review and approved by the IRB at Universidad San Francisco de Quito (2020-023M). Information from patients was anonymized before analysis. Patients offered their oral consent for gathering demographic data. Health care personnel followed intra-hospital guidelines to fill information forms considering their home addresses according to HGSQ policies.

### Transitioning to a COVID-19 reference hospital

From March 10^th^ to March 28^th^, 2020, the departments of infectology and epidemiology of the HGSQ, in coordination with others services, developed the ‘Virus in Movement (VM)’ protocol for the COVID-19 crisis management; which encompassed the following: every time that a patient suspected of COVID-19 needed transit to the imagenology department—or other hospital setting—the VM alert was immediately activated and triggered three coordinated steps: (a) evacuation of all health personnel and patients from the movement areas (e.g., halls, imagenology department, etc), (b) blocking all doors to avoid transit of other persons, and (c) usage of an exclusive elevator. After movement, areas occupied by suspected/positive patients were cleaned with pulsed-xenon ultraviolet room disinfection according to Jinadatha et al. (2015) and Kovach et al. (2017) [27,28].

### AI-aided assisted Chest computed tomography (CT) screening for patient’s triage

Considering the limiting testing capacities and the over-the-edge public health system in a developing country such as Ecuador, the implementation of a novel triage strategy was paramount [13,14,19]. We used non-contrast chest CT scans to stratify COVID-19 suspected patients according to a severity score calculated via AI for assisted COVID-19 CT screening. The software was developed as part of the Huawei Cloud AI services [29], implemented for the first time in Latin America for the HGSQ imagenology department [30]. The Huawei Cloud AI-assisted CT diagnosis software can be described as a deep-learning neuronal network approach for automated medical image segmentation for identification of abnormalities on chest CTs [31,32], it was released on March 17th, and uses MindSpore as its AI deep-learning algorithm framework, which was developed entirely by Huawei [33,34]. For calibration purposes, the AI has been trained with ∼4,000 chest CT images from confirmed positive COVID-19 cases from China [29]. Scanned CT images were uploaded to the hospital picture archiving and communication system (PAC) and then examined with the Huawei AI to detect the presence GGOs and lung consolidations [29,35].

By calculating internal metrics comparing the predicted lesions from the trained AI, with the actual lesions from the CT scan [35], the AI-aided assisted CT screening provided a score that categorized patients in three classes: non-severe (score of 0-30%), moderately severe (30-70%), and severe (>70%); considering the likelihood of being COVID-19 positive in relation with the severity of radiological findings on chest CT [29,36,37]; a medical radiologist examined and confirmed severity scores. Depending on this categorization, patients were distributed in different sections of the hospital (i.e., score rooms) to prevent the spread of the virus. Similar AI-based approaches for imaging recognition on chest CTs have been deployed in China with contrasting results for COVID-19 diagnosis and the majority of them still required further evaluation [19,38,39].

### Clinical and epidemiological characteristics of COVID-19 patients

Epidemiological (e.g., hospitalization time, risk factors, source of infection), clinical (e.g., symptoms and signs), and laboratory data, together with drug treatment schemes, were recovered from medical records of hospitalized patients with respiratory symptoms above 18 years old with laboratory confirmation of SARS-CoV-2 via RT-PCR. We considered data from patients either death or discharged admitted in the HGSQ during February 29^th^ to March 28^th^.

### Spatial distribution of households of patients and health personnel

Due to the risk of health workers spreading COVID-19 to other hospitalized patients or the community in the context of asymptomatic SARS-CoV-2 carriers, and the lack of reliable testing for antibody detection [25,26,40], all health personnel attending inpatients with presumptive or confirmed diagnosis of COVID-19 at the HGSQ completed online forms disclosing attending time, home address, and use of personal protective equipment (PPE). Health workers with symptoms related to COVID-19 were treated by the department of occupational medicine and immediately notified to the infectology and epidemiology department to suspend their activities during 14 days after symptom resolution and RT-PCR negative tests as suggested by different guidelines [20,21,41]. We used the information on these forms to suggest potential clusters of COVID-19 monitoring outside hospital settings by georeferencing addresses of health workers using Google Maps (https://www.google.com/maps/), calculating the distance to the hospital (i.e., HGSQ), and estimating the amount of people at risk of infection considering the population density at three distance buffers centered at the hospital using the 2010-2020 population projections from the official Ecuadorian census (https://www.ecuadorencifras.gob.ec/proyecciones-poblacionales/). Distance calculation and population at risk was calculated using TerrSet (version 18.39; https://clarklabs.org/terrset/). Coordinates and results of this analysis were plotted in maps using QGIS (3.4 Madeira; https://qgis.org/es/site/forusers/download).

## Results

### Transition to COVID-19 hospital management center

Considering the impending pace of the epidemic, we had to implement our strategy fast. On March 13^th^, an outside hospital triage area was established to receive the first COVID-19 case in the HGSQ (Fig. 2). At that moment, all services (e.g., neurology, surgery, gynecology, etc) were attending patients with other pathologies. In order to prevent intrahospital transmissions, different actions were implemented as follows. During the first 24 hours of the first COVID-19 case admission, the VM protocol was activated four times (i.e., four VM events). The average duration of the implementation of the full VM protocol was about 1 hour per event. A total of 75 inpatients were transferred to different portions of the hospital a total of 432 times under the VM protocol in a period of 18 days of transition. No health care worker or other person transiting within the hospital got infected with SARS-CoV-2 in this period.

**Figure 2.**
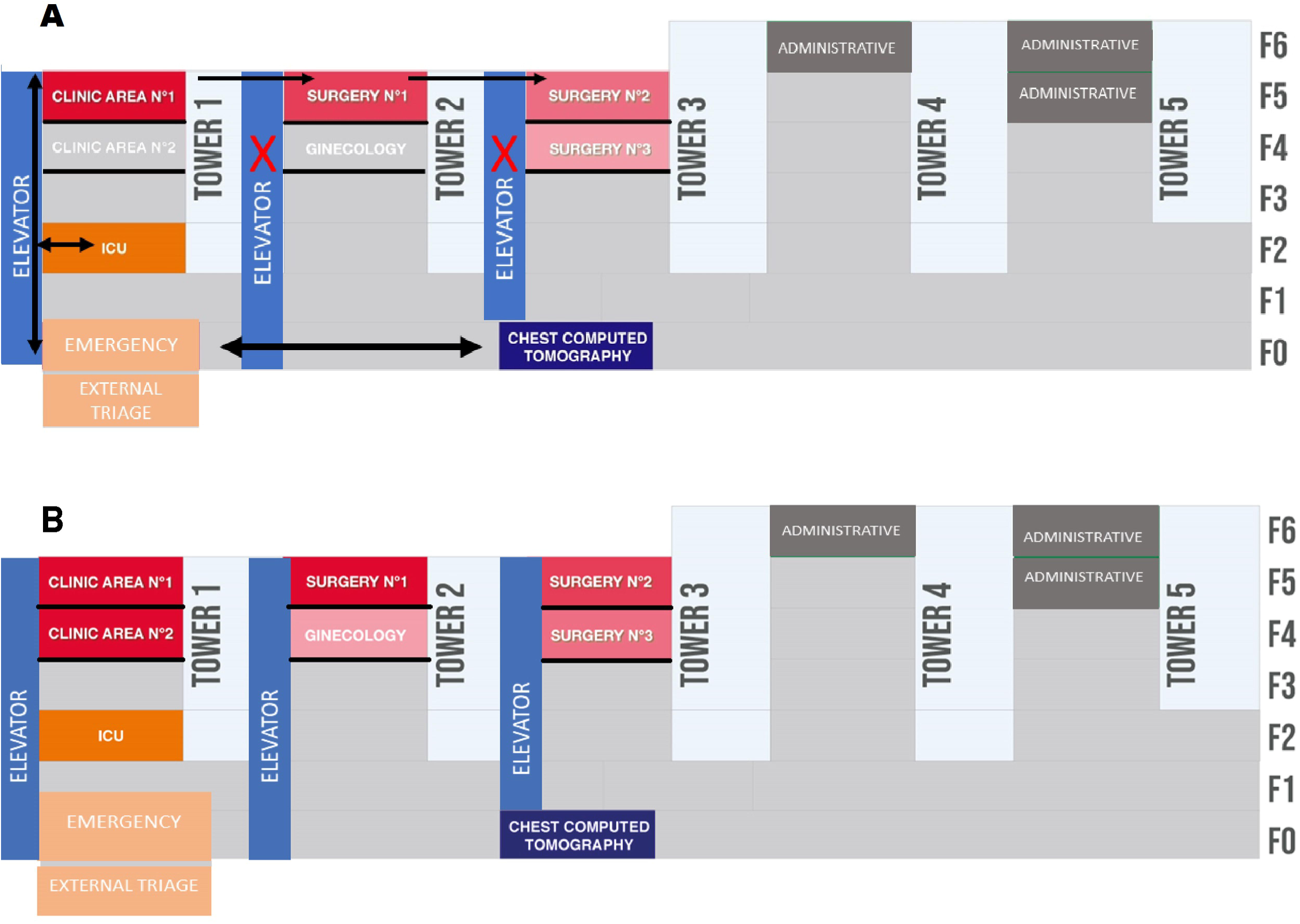
Schematic representation of COVID-19 exclusive rooms (score rooms in red) established during hospital transition. The initial distribution of score rooms (A) was complemented with newer areas to organize confirmed, suspected, and negative patients based on laboratory diagnosis and computer tomography assisted severity score (B; see text). Arrows (black) are the same for panels A and B but are depicted only for A, they summarize the available flow of beds around the hospital with only one elevator available (golden vertical arrow to the left); other elevators were disabled (red cross) in order to fully control movement of personnel. The different floors of the hospital are labeled as F0-F6 (right).

The increasing incidence of suspected COVID-19 patients forced a redistribution of hospital areas which, due to the lack of timely RT-PCR testing, was guided mainly by the AI assisted chest CT scan scores. The services of infectology and epidemiology designated ‘score rooms’ as exclusive rooms for different COVID-19 patient categories. Specifically, cases with a higher probability of COVID-19 (score >70%) were hosted on the clinic area N°1, patients with moderate probability (score 30-70%) were concentrated on surgery area N°1. Lastly, patients with a score less than 30%, and therefore a lower likelihood of COVID-19 positivity, were located in surgery area N°2 and N°3 in common rooms separated by gender (Fig. 2). We prevented the CT machine to act as a fomite source of SARS-CoV-2 transmission, using disinfection based on conventional cleaning followed by pulsed-xenon ultraviolet room sterilization [27,28].

A further reorganization was put in place once confirmatory RT-PCR tests became available, although AI-assisted chest CT scans remained the main approach driving triage considering the lag of laboratory-based testing results, and their application only to selected patients (i.e., with higher suspicion of infection). Molecularly confirmed SARS-CoV-2 patients were located in clinic area N°1 now exclusive for these patients. Surgery area N°1 became exclusive for patients with a score above 70%, surgery area N°2 became exclusive for patents with a 30-70% score, and clinic area N°2 was suited for those patients with a score less than 30%. Thus, clinic areas N°1 and N°2, together with surgery areas N°1 and N°2 became SARS-CoV-2 exclusive wards for laboratory confirmed diagnosis together with those with a CT-score >70%. Moreover, other areas of the hospital were accommodated to host patients with a moderate likelihood of COVID-19 infection (i.e., moderate AI-CT score) including the gynecology floor, for patients with an score between 30-70% and the surgery area N°3, divided in two sections: one for patients with an score less than 30% and the other for negative RT-PCR patients in the path of discharge.

### Efficacy of chest CTs aided by AI for COVID-19 triage

We obtained chest CTs for 75 patients with laboratory confirmed SARS-CoV-2 diagnosis (Table 1). Images showed that the distribution of the GGO in the lungs were most peripheral (30/61, 49.18%) than central, the latter present in 21 of the 61 positive cases (34.43%). Bilateral lesions were predominant. Five laboratory positive patients showed an absence of GGO patterns in the lungs (8.2%). Seven SARS-CoV-2 negative patients showed peripheral GGO lesions (7/14, 50%), while four showed central GGOs (4/14, 28.57%).

**Table 1.** General characteristics, risk factors, and symptoms/signs.

|  | Positive (%) | Negative (%) | Totals (%) |
| --- | --- | --- | --- |
| RT-PCR confirmation | 61 (81.3) | 14 (18.6) | 75 (100) |
| Median age (range) | 50 (25-76) | 49 (27-77) | 50 (25-77) |
| Male | 39 (63.9) | 8 (57.1) | 47 (62.6) |
| Female | 22 (36) | 6 (42.3) | 28 (37.3) |
| ICU | 14 (23) | 4 (28.6) | 18 (24) |
| • ICU male | 11 (18) | 2 (14.3) | 13 (17.3) |
| • ICU female | 2 (3.3) | 3 (21.4) | 5 (6.6) |
| Non-ICU | 47 (77) | 10 (71.4) | 57 (76) |
| • Non-ICU male | 28 (45.9) | 6 (42.9) | 34 (45.3) |
| • Non-ICU female | 19 (31.1) | 4 (28.6) | 23 (30.6) |
| Risk factors |  |  |  |
| Contact with European tourist | 3 (4.9) | 1 (7.1) | 4 (5.3) |
| Contact with COVID-19 case | 10 (16.4) | 4 (42.9) | 14 (18.6) |
| Unknown contact | 14 (23) | 6 (42.9) | 20 (26.6) |
| Travel to Guayaquil | 26 (42.6) | 3 (21.4) | 29 (38.6) |
| Travel to other Ecuadorian cities | 5 (8.2) | 0 (0) | 5 (6.6) |
| Travel to European | 2 (3.3) | 0 (0) | 2 (2.6) |
| countries |  |  |  |
| Health worker | 1 (1.6) | 0 (0) | 1 (1.3) |
| Symptoms/signs |  |  |  |
| Fever | 59 (96.7) | 10 (71.4) | 69 (92) |
| Cough | 48 (78.7) | 13 (92.9) | 61 (81.3) |
| Odynophagia | 36 (54) | 5 (35.7) | 41 (54.7) |
| Headache | 13 (21.3) | 2 (14.3) | 15 (20) |
| Diarrhea | 12 (19.7) | 1 (7.1) | 13 (17.3) |
| Vomiting | 5 (8.2) | 0 (0) | 5 (6.7) |
| Anosmia | 4 (6.6) | 1 (7.1) | 5 (6.7) |

For 75 COVID-19 cases categorized as positive/negative by laboratory confirmation from March 10th to March 28th. Percentages from the first row are calculated in relation to the total 75 cases. Percentages of the following rows are calculated in relation to the amount of positives, negatives, and totals in the first row.

Sensitivity and specificity indexes were calculated based on the ability of the Huawei AI-assisted CT screening system to correctly identify cases as COVID-19 confirmed by RT-PCR, whenever they were categorized with a higher score (i.e., >70%). We obtained severity scores for 37 laboratory-tested patients (49.3%). Sensitivity corresponded to 20.7% and specificity to 66.7% when considering the likelihood to classify a patient as COVID-19 positive with a score over 70%. Thus, 7/28 positive and 3/9 negative patients (n =10) were allocated in 70% score rooms, 10/20 positive and 1/9 negative patients (n = 11) in score rooms for 30-70% score, and 11/28 positive and 5/9 negative patients (n = 16) were allocated in rooms for scores less 30%.

### Epidemiological characteristics and description of clinical cases

At the moment of data collection, we have attended 2,590 patients with respiratory symptoms, 93 positive to SARS-CoV-2 (i.e., 3.5% prevalence). We are still waiting for laboratory confirmation of 90 patients. We have discharged patients with two RT-PCR negatives and recorded a total of 18 deaths. We present clinical, epidemiological, and laboratory data, together with treatment schemes for 61 SARS-CoV-2 positive cases and 14 negatives, but COVID-19 suspected (n = 75; Tables 1-4). The mean age of patients from both groups is 50 years old with a male majority (male/female ratio: 47/28 = 1.67; Table 1).

From the positive patients, 42.6% (n = 26) reported having traveled to Guayaquil, the city with more cases of COVID-19 in Ecuador (Ministerio de Salud Pública del Ecuador 2020; Fig 1, Supplementary_1). Moreover, 16.4% (n = 10) positive cases reported to have a history of close contact with known COVID-19 patients (Table 1). In general, fever, cough, and odynophagia were the most prevalent symptoms while anosmia was the least common (Table 1). At least ten negative patients also had fever (71.4%) and 13 presented cough (92.9%); one of the negative cases also referred anosmia (7.1%). The average number of days from onset of respiratory symptomatology until hospital attention was eight days (ranging from zero to 20 days).

All cases admitted to the intensive care unit (ICU) presented values of C-reactive protein(CRP) above 10 mg/L and LDH above 250 UI/L. Median values from non-ICU patients also presented higher values for LDH and borderline values of CRP (Table 2). From laboratory positive SARS-CoV-2 cases, those admitted at the ICU presented higher values of D-dimer than those outside, but in both classes the median was higher than normal. The median value for levels of transaminases AST and ALT were slightly altered for ICU patients (Table 2). Procalcitonin was altered in patients admitted to the ICU with lesser values on patients attended outside this unit. Creatinine was within normal ranges for both ICU and non-ICU cases (Table 2). One case at ICU presented *Candida* spp. in trachea, other patients in ICU with normal values of procalcitonin presented *Klebsiella pneumoniae* and *Pseudomonas aeruginosa*-antibiotic-sensitive in blood, and *Candida spp* in urine. Leucocytes and platelet counts for both case categories were within normal values, but lymphocytes were lower for ICU patients (Table 3).

**Table 2.**
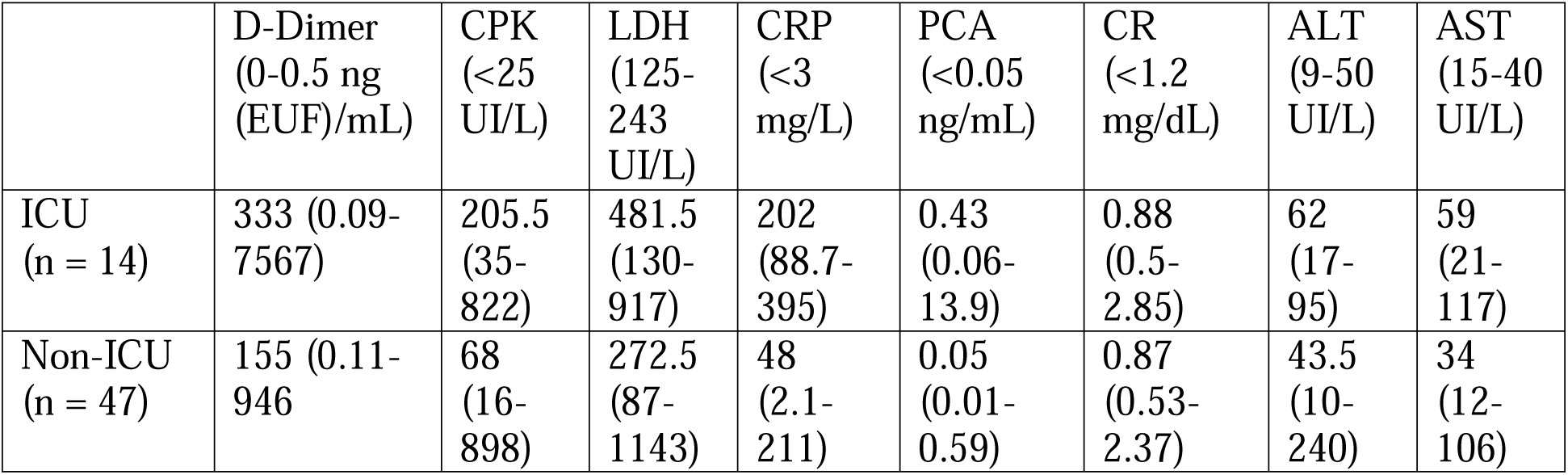
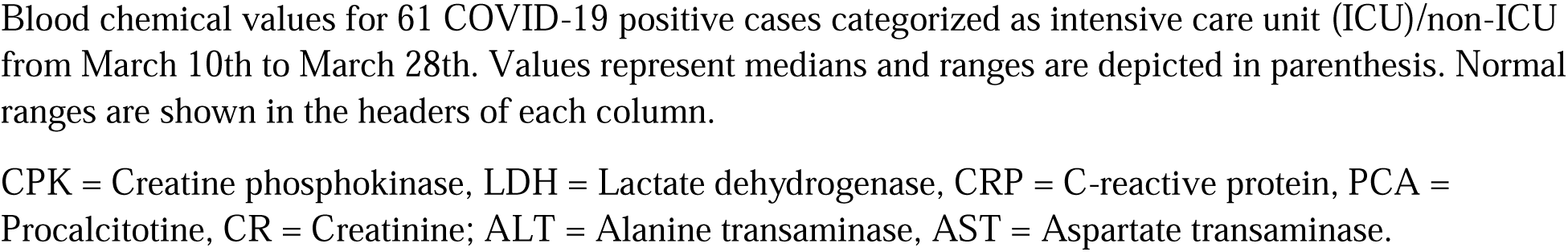
Blood chemical values

**Table 3.** Biometry results.

|  | Leucocytes<br>(3500-9500/ml) | Lymphocytes<br>(1100-3200/ml) | Lymphocytes<br>(20-40%) | Platelets<br>count<br>(125000-350000/ml) |
| --- | --- | --- | --- | --- |
| ICU<br>(n = 14) | 8900<br>(4800-15000) | 1000 (500-1800) | 11 (4.2-21) | 183000<br>(145000-277000) |
| Non-ICU<br>(n = 47) | 5700<br>(1900-13300) | 1440 (500-2490) | 22.5 (8.9-61.7) | 210000<br>(114000-452000) |

Leucocyte, lymphocyte, and platelet counts for 61 COVID-19 positive cases categorized as intensive care unit (ICU)/non-ICU from March 10th to March 28th. Values represent medians and ranges are depicted in parenthesis. Normal ranges are shown in the headers of each column.

Only two negative SARS-CoV-2 patients received treatments without chloroquine (Table 4). Most schemes were associated with beta-lactam antibiotics such as ceftriaxone, piperacillin/tazobactam, or meropenem (Table 4). A total of 45/75 patients were discharged. The average stay in hospital was of 10 days (range = 8-20 days); these patients received the scheme based on ceftriaxone, azithromycin, and chloroquine for 7 days (Table 4).

**Table 4.**
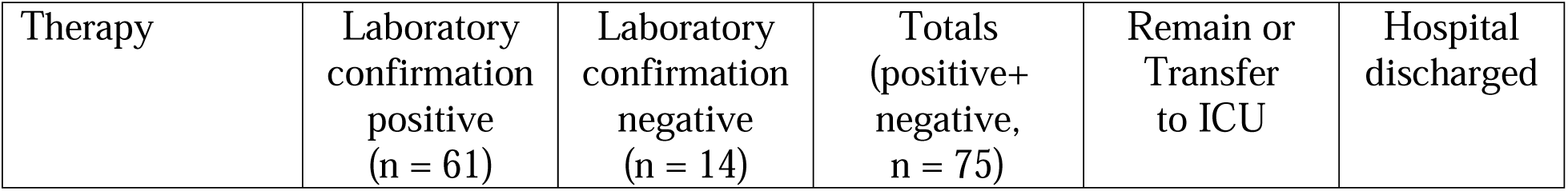
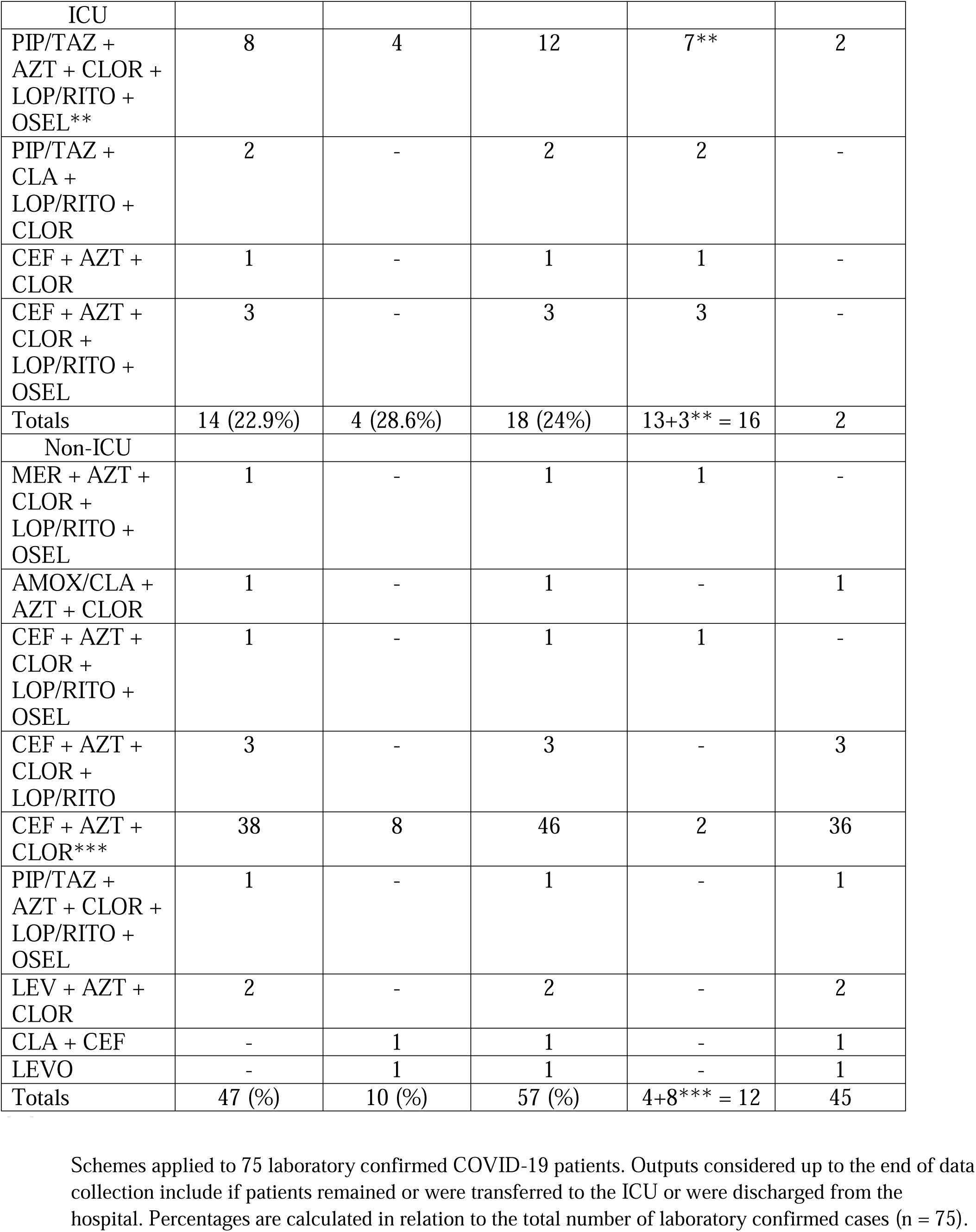
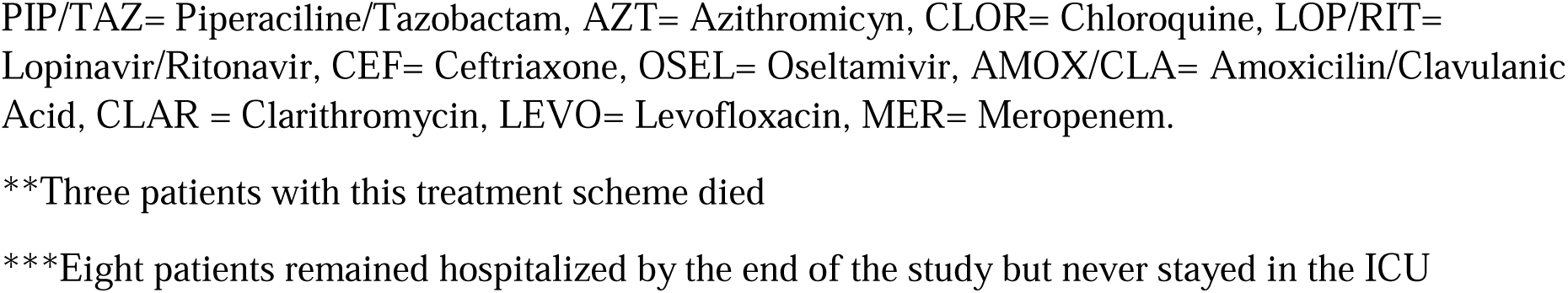
Treatment schemes

### Health personal contact trace and georeferencing

The first COVID-19 case had contact with at least 51 health workers. Our first ten patients had contact with a mean of 19 health workers (range = 9-51). After the transition to a COVID-19 exclusive hospital, we reduced contact with health personnel to a mean of five (range = 3-7). By georeferencing domiciles of health workers, we identified than 43 live between 0 to <5 km from the hospital in an urban area with 783,291 inhabitants. Similarly, 39 health workers lived around 5 to <10 km from the hospital in an area with 665,928 inhabitants. Finally, 22-health workers lived between 10 to <15 km around the hospital in an area with 558,639 inhabitants. The majority of health workers came from the south and northernmost metropolitan area. By this analysis, we also identified that some health workers (n = 4) travel up to 85 km to arrive to the HGSQ, crossing other provinces including Imbabura to the north and Cotopaxi to the south (Fig. 3).

**Figure 3.**
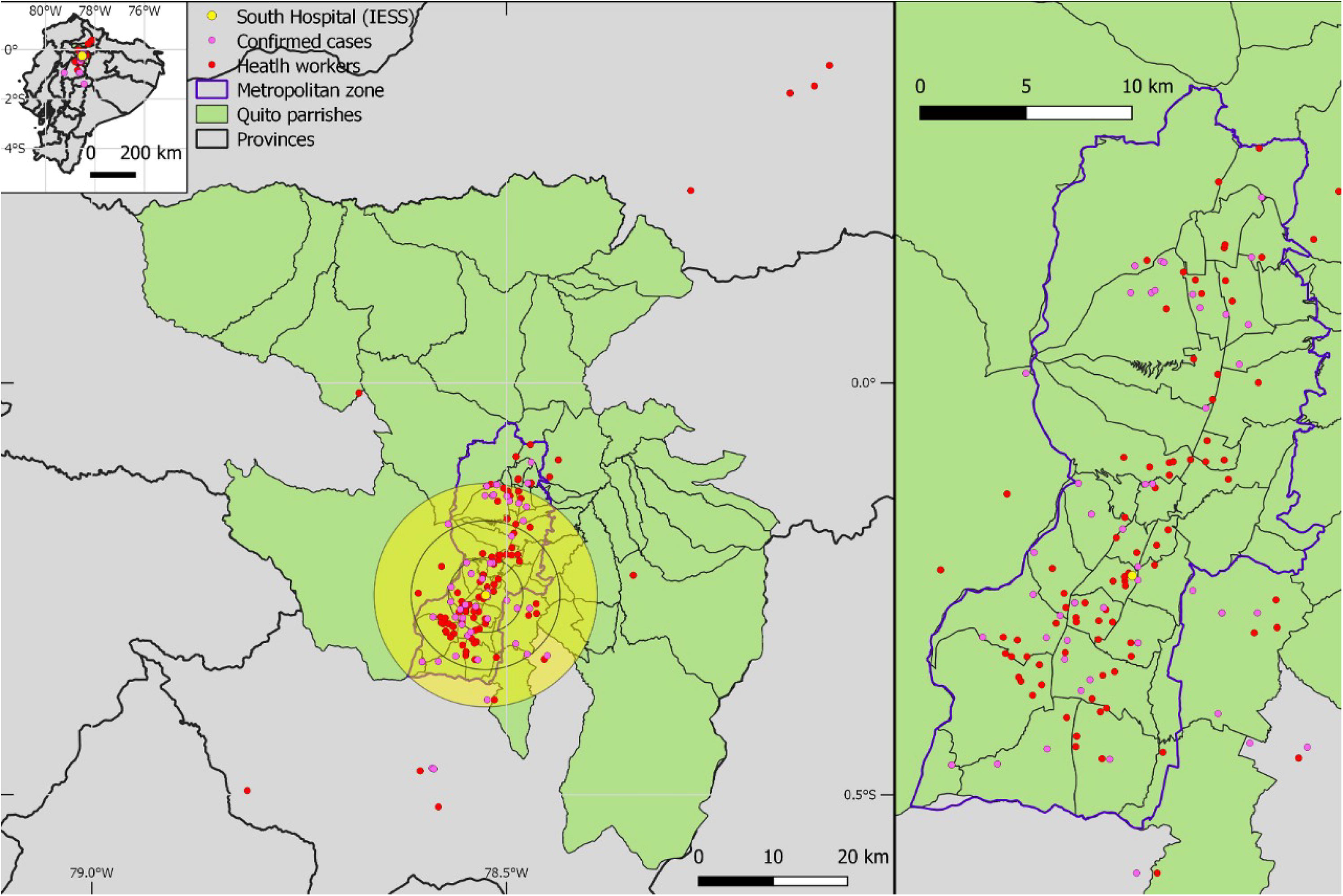
Distribution of 54 COVID-19 cases and 126 health workers from the Hospital General del Sur de Quito (HGSQ), in Quito, Ecuador.

Domicile georeferencing was completed for 54 out of 75 (72%) patients with COVID-19 positive diagnosis. At the beginning of the epidemic, the majority of patients came from the south of Quito city. The geographic distribution of cases changed once the hospital became exclusive for COVID-19, with cases broadened to encompass the whole Quito metropolitan area and its surroundings including one case arriving from the southern province of Cotopaxi (Fig. 3).

## Discussion

Ecuador faced the current COVID-19 pandemic following WHO recommendations [42], however, despite being supported by the central government, main cities managed their health crisis differently. Despite the National Emergency Operations Committee (COE, Spanish) recommended social isolation and to stop all mobility on March 16^th^ [6], Before the pandemic, Guayaquil had a sub-employed working population of 16.2% [43] with urbanistic features (e.g. slums and lack of water provision) that complicate the implementation of social isolation measures [44]. Quito stopped all face-to-face academic activities earlier than the COE recommendations (March 12^th^; [45,46]), and followed social isolation with constrained mobility and mass gatherings strictly, buying the HGSQ time to develop the aforementioned approaches, which, among others, effectively allowed us to transit to a COVID-19 exclusive attention center. In our hospital, strategies of management, including change in distribution of suspected COVID-19 patients has allowed us to halt the high incidence of nosocomial infections reported elsewhere [17] which remained in zero until data collection (March 28^th^).

The inclusion of the Huawei Cloud AI-assisted CT screening for COVID-19 in the HGSQ represent the first attempt to explore the ability of this tool to aid different hospitals, which received the software for free [37]. The same tool has been deployed for hospitals in other countries including China, Malaysia, and the Philippines [47,48]. For our particular case, the tool was used for initial triage of suspected patients due to the lag between RT-PCR testing and results availability (range 28-120 hours), concentrating all efforts to stop a potential COVID-19 nosocomial outbreak. However, even for the most severe cases, with a higher likelihood of COVID-19 infection, we obtained unacceptable test accuracy values for sensitivity and specificity (i.e., 20.7% and 66.7% respectively); this has two immediate consequences. First, from a research perspective, the performance of the AI-assisted CT screening for COVID-19 using Huawei technology is poor enough to actually recommend it; something that has also been adverted by the Philippine College of Radiology□[49]. Second, from a pragmatic perspective, the HGQS continue to rely on this method for patient triage since at least relieve one of the bottlenecks of attention in the context of the current burgeoning epidemic: patients can be allocated to specific areas depending on the severity score to prevent COVID-19 spread, a crucial endeavor considering the potential correlation between increasing health burden and mortality [50].

In order to fully test the ability of this AI-based screening system, a strict study design should evaluate both laboratory confirmed positive and negative cases [15,18,19,51]. We currently lack information from the latter since it is impossible to have the luxury of using the only CT facilities of the hospital to expose non-suspected patients to infection. Nevertheless, during the development of this study, zero cases have been associated with exposition of patients or health workers to the CT area, which shows preliminary evidence that the routine cleaning plus pulsed-xenon ultraviolet disinfection approaches effectively prevent the CT to become a fomite source of SARS-CoV-2 contagion; [27,28]. It is important to notice that the Huawei Cloud AI-assisted CT screening software was donated freely to the HGSQ and we took the advantage to scientifically evaluate it. Currently, there are many other AI-medical image processors that have been deployed, especially in China, and that merit further assessment as well [38].

Patients attended here were part of the first COVID-19 outbreak in Quito. Despite our limited sample of laboratory confirmed positive cases (n = 61), demographic and clinical characteristics of COVID-19 infection were similar to that of previous reports [17,52,53], namely, a majority of male individuals with a median age of 50 years presenting fever, cough, and odynophagia (Table 1). Although less frequent, we also found cases describing anosmia, which has been correlated with positive COVID-19 infections [54]. In the present study we did not find cutaneous manifestations of the disease [55]. Blood chemical markers such as CRP, LDH, CPK, etc were increased in patients admitted to ICU in comparison with non-ICU patients (Table 2). Values for D-dimer, were above normality for ICU and non-ICU patients but higher for the former than the latter (Table 2); recently, coagulopathies have been incriminated as drivers of mortality to COVID-19 positive patients [17,56]. From our variables considered for blood count, only lymphopenia was apparent for ICU cases (Table 3).

In Ecuador, RT-PCR laboratory testing for SARS-CoV-2 was centralized in few hospitals, institutes, and private laboratories, up to 46 days since the beginning of the epidemic [12]. This factor influenced diagnosis delay, epidemic spread, and the urgency to implement out-of-the-box triage approaches such as the one presented here [13,14,57]. By the time of this study, testing was limited to 7.46 per 10,000 inhabitants [4], thus, clinical suspicion of patients with respiratory symptoms with elevation of CRP, LDH, and lymphopenia (Table 2 and 3), can be useful markers for triage in settings unable to rely on molecular or radiological tests [13,14].

In this study, the majority of laboratory confirmed cases received treatment schemes based on the combination of chloroquine plus azithromycin except for two negative cases, which received clarithromicine plus ceftriaxone or levofloxacin (Table 4). Gautret et al. (2020), showed that the treatment based in hydroxychloroquine plus azithromycin was associated with viral load reduction/disappearance in a small sample size of COVID-19 patients [58]. Two recent clinical trials (currently on pre-print stage), assessing the safety and efficacy of hydroxychloroquine and chloroquine recommended to avoid COVID-19 treatment schemes with any of these drugs due to the higher mortality detected [59,60]. Regardless, a more recent study showed a lack of association between treatments including hydroxychloroquine and development of poor clinical outcomes [61]. We were unable to investigate electrocardiogram QT alterations as previously reported [62]. Due to the lack of control groups, our findings should be interpreted as preliminary and by no means as evidence to support any treatment scheme, which is still a topic largely debated with no consensus [21]. Lopinavir and Ritonavir treatment was exclusively used in ICU patients, a case control study published on March 18^th^ suggested a lack of effect in death reduction [63,64], however ICU attendant from this hospital decided to continue with the antiviral treatment scheme due to the lack of literature consensus and an apparent clinical improvement still on quantification.

Our spatial analysis depicted a spatial proxy of population at risk of infection considering the presence of potential carriers [25]. During the progression of the epidemic in Ecuador, officers from the Ministry of Public Health have referred to ∼1,500 health workers getting infected with SARS-CoV-2 while minimizing the need of full body personal protection [65]. We remark the need to protect health workers and provide them adequate personal protective equipment; if this requirement is unmet, a large susceptible population might be at risk of infection both in the hospital and the community [66] (Fig. 3).

It is important to remark that the case of the HGSQ is uncommon in comparison to other Ecuadorian health facilities. Health sector in Ecuador is fractionated encompassing public, social security, military, police, and private health providers [5]; thus, hospital management and protocols are far from standardized [67]. The information published here might aid the implementation of protocols in other regions of Ecuador and also other regions of Latin America, where overrun health systems aimed to control the current epidemic, or potentially future emergent respiratory transmitted diseases, are in need of local perspectives [68].

## Data Availability

All Data is available

## Acknowledgments

We do not express any particular acknowledgment.

